# Etiological spectrum of non-compressive myelopathies in Uganda: a prospective, observational study using neuroimaging, autoantibodies, and next-generation sequencing

**DOI:** 10.1101/2025.09.03.25335017

**Authors:** Abdu K Musubire, Kristoffer E Leon, Eoin P Flanagan, Prashanth S Ramachandran, Chloe Gerungan, Kelsey C Zorn, Annie Wapniarski, David B Meya, Paul Mubiri M., Twaha Kisozi, Kimbugwe Denis, Mahsa Abassi, David R Boulware, Vyanka Redenbaugh, Jessica Sagen, Joseph L DeRisi, Cynthia T Chin, Paul R Bohjanen, Patrick Cras, Barbara Willekens, Sean J. Pittock, Michael R. Wilson

## Abstract

**Background:** Non-compressive myelopathy can lead to severe disability and death. The diagnosis is often complex and resource-intensive, with regional variation in causes. Data on the epidemiology and etiology of non-compressive myelopathy in Africa remains limited.

**Methods:** A prospective observational study of adults presenting with clinical signs and symptoms of non-compressive myelopathy was conducted. Patients were recruited from 2013-2015 and 2018-2022 in Kampala, Uganda. Participants underwent spinal magnetic resonance imaging (MRI) to exclude extradural spinal cord lesions. Serum and cerebrospinal fluid (CSF) were tested for aquaporin-4 (AQP4) and myelin oligodendrocyte glycoprotein (MOG) antibodies. Metagenomic next-generation sequencing (mNGS) was performed on CSF to detect infectious etiologies. Serum vitamin B12 levels and tissue biopsies were performed at clinician discretion. Participant characteristics and diagnostic findings were summarized using descriptive statistics.

**Findings:** Among 420 participants screened, 144 were enrolled and included in this analysis. The median age was 33 years (interquartile range 25-44). 79 participants (55%) were male, and 38 (26%) were living with HIV. Intradural abnormalities were identified on spinal MRI in 79 (55%) participants. An etiologic diagnosis was established in 50 (35%) participants, including neuromyelitis optica spectrum disorder (NMOSD, n=10), myelin oligodendrocyte glycoprotein antibody-associated disease (MOGAD, n=7), *Schistosoma mansoni* (n=9), cytomegalovirus (n=1), varicella zoster virus (n=1), vitamin B12 myelopathy (n=8), spinal cord tumor (n=12), and arteriovenous malformation (n=2).

**Interpretation:** This study identified a wide range of etiologies for non-compressive myelopathy in Uganda. Autoimmune myelopathies (NMOSD and MOGAD) accounted for 14% of cases, while mNGS identified an infectious cause in 13% of participants. These findings highlight the importance of expanding access to both autoimmune and infectious diagnostic testing in resource-limited settings.

**Funding:** This study was supported by the NIH (R01NS113828, R01AI145437, R25TW009345, K24AI096925, and 1K43TW010718) and the Westridge Foundation (MRW). Sequencing was performed at the UCSF Center for Advanced Technology, supported by UCSF PBBR, RRP IMIA, and NIH 1S10OD028511-01 grants. The K24AI096925 was used in the design of the study and data collection, R25TW009345 was used in data collection while 1K43TW010718 was used in analysis, interpretation of the data and in writing the manuscript.

## Introduction

Non-compressive myelopathy is defined as a non-traumatic spinal cord injury without detectable radiological evidence of spinal cord compression.^1^ Non-traumatic myelopathies can cause significant disability or death, posing a considerable physical and economic burden.^2^ The prevalence of non-traumatic myelopathy varies geographically, with reported rates of 112 cases per 100,000 people in Canada and 231 cases per 100,000 people in India.^3^ Incidence and prevalence of non-traumatic myelopathy in medium- and low-income countries, while not well documented, are generally believed to be higher and affecting a younger population than those in high-income countries.^3,4^

The causes of non-compressive myelopathy include autoimmune, infectious, metabolic, vascular, neoplastic, and idiopathic factors. Considerable geographical variation exists with autoimmune causes, such as multiple sclerosis (MS), predominating in temperate, high-income regions, whereas infectious causes are more common in tropical regions, primarily in middle- and low-income countries.^5–7^ Limited data on autoimmune causes of non-compressive myelopathy likely reflect restricted access to diagnostic testing. Population-based studies in developed countries show that autoantibody testing for aquaporin-4(AQP4-IgG) and myelin oligodendrocyte glycoprotein antibody (MOG-IgG) has reclassified approximately 12–14% of patients previously diagnosed with idiopathic transverse myelitis (TM) as having neuromyelitis optica spectrum disorder (NMOSD) or MOG antibody-associated disease (MOGAD).^4,8–11^ This is consistent with the pattern previously observed in hospital-based reports of MS from Kenya and Zambia.^12,13^ Recent advances in diagnostic tools for both autoimmune and infectious etiologies have significantly improved the ability to identify the underlying causes of non-traumatic myelopathy and presents an opportunity to address this diagnostic gap.^4,8^ This is important because early diagnosis allows treatment with specific therapies.

Epidemiological data on non-compressive myelopathy across Africa is significantly lacking. To address this gap, we conducted a prospective cohort study to assess for the different etiologies of non-compressive myelopathy in Uganda, utilizing a comprehensive diagnostic approach that included magnetic resonance imaging (MRI), antibody testing for AQP4-IgG and myelin MOG-IgG in serum and cerebrospinal fluid (CSF), and metagenomic next-generation sequencing (mNGS) of DNA and RNA extracted from CSF for unbiased infectious disease testing.

## Methodology

### Study design and participants

Participants with non-traumatic spinal cord injury from Mulago National Referral Hospital from 2013 to 2015 and Kiruddu National Referral Hospital from 2018 to 2022, in Kampala, Uganda were prospectively enrolled.^14^ Ugandan adults, who presented with motor (paraplegia or quadriplegia with upper motor neuron signs or features consistent with spinal shock), sensory (sensory level for pinprick and light touch and/or loss of proprioception and vibration), and/or autonomic dysfunction (impaired sphincter control), were screened for participation into the study. All study participants underwent an MRI of varying spinal cord segments dependent on the clinical scenario and neuroanatomic localization. Lumbar puncture was performed within three days of study enrollment. After hospital discharge, study participants were followed every three months for one year to assess clinical status and vital outcomes.

The Makerere University Research and Ethics Committee (REF 2013-056) and the Uganda National Council for Science provided regulatory approval for the study. All participants provided written informed consent to participate in the study.

### Study Procedures

Baseline demographic and clinical data were collected at the time of presentation by study personnel. A comprehensive neurological examination was performed at presentation by the study physician. The severity of the spinal cord injury was determined using the American Spinal Injury Association (ASIA) impairment scale.^15^

Depending on the neurological localization of the spinal cord lesion, focused MRI of the spine (cervical, thoracic, and/or lumbar) was obtained at baseline. MRI was performed on a 16-channel 1.5 T MRI (Philips Achieva 1.5 T, Phillips Healthcare, the Netherlands) at the Kampala MRI Center. T1, T2, fluid attenuated inversion recovery (FLAIR), and short tau inversion recovery (STIR)-weighted MRI images were obtained. Post-gadolinium images were not obtained systematically. Spinal cord lesions were categorized as either compressive or non-compressive. Non-compressive myelopathy cases were further categorized as either imaging negative (i.e., no abnormalities seen on the spinal cord MRI) or as abnormal imaging, including lesions suggestive for tumor, inflammation, vascular abnormality, metabolic deficiency or not classifiable. A longitudinally extensive spinal cord lesion, termed longitudinally extensive TM (LETM) when an inflammatory cause is presumed or confirmed, was defined by a spinal cord lesion extending across three or more vertebral segments. Short transverse myelitis (TM) was defined as an inflammatory myelopathy with a lesion spanning fewer than three vertebral segments. Two independent physicians, blinded to all other diagnostic test results, reviewed and adjudicated MRI images: a neurologist with experience in neuroimmune disorders (BW) and a board certified neuroradiologist (CTC). Disagreements were addressed through consensus or, when necessary, reviewed by a third party to reach consensus.

Study participants underwent blood and CSF collection at the time of study enrollment. CSF was collected in three separate tubes: tube one was sent for routine testing, including cell count, glucose, protein, Gram stain, and bacterial culture; tube two was placed in RNA-stabilizing media for mNGS; and tube three was centrifuged, with both supernatant and cell pellet stored in 0.5-1.0 mL aliquots at -80⁰C. Serum vitamin B12 levels were measured using a chemiluminescent immunoassay on a Cobas Integra analyzer. Biopsies were performed at the discretion of the treating physician.

### CSF Metagenomic Next-generation Sequencing

Stored CSF samples were sent to University of California, San Francisco (UCSF) for mNGS. A detailed description of mNGS methods are described by Ramachandran et al.^16^ In brief, total nucleic acid was extracted from 90 µL of CSF using the Zymo Quick-DNA/RNA MagBead kit. The nucleic acid was then divided, with half subjected to DNAse treatment to isolate RNA and the remainder used for DNA sequencing (DNA-Seq). Total nucleic acid was also extracted from water samples as a control. RNA sequencing (RNA-Seq) and DNA-Seq libraries were prepared in accordance with the protocol using New England Biolabs’ NEBNext Ultra II RNA and DNA library preparation kits respectively. Bulk library preparation was carried out with the Echo Labcyte 525 and Agilent Bravo or Integra Viaflo 96 liquid handling robots. Host ribosomal RNA was removed using the Qiagen QIAseq Fast Select RNA removal kit. Pooled libraries were then size selected with Ampure beads before being sequenced on an Illumina Novaseq 6000 using 146 base pair paired-end sequencing.

mNGS data were analyzed using CZ ID (Chan Zuckerberg ID), an open-source, cloud-based metagenomics platform for microbial detection and pathogen discovery^17^. The pipeline’s first step involves aligning the raw sequences to the human genome and removing human aligning sequences. Subsequent steps in the CZID pipeline include the removal of low-quality reads, duplicate reads, and additional human sequence filtering, further reducing contaminating and uninformative sequences from the nonhuman dataset. The remaining sequences are assembled and aligned using an indexed version of the NCBI’s GenBank database to determine the source of non-human sequences in the datasets. Criteria for determining an infection are reported in the Supplemental Methods and also included clinical adjudication.

### Autoantibody Testing

Stored paired serum and CSF samples were sent to The Mayo Clinic for AQP4-IgG and MOG-IgG testing using a live-cell-based flow cytometry assay with fluorescence-activated cell sorting. Samples were heated at 56°C for 35 minutes to inactivate complement proteins. For AQP4-IgG detection, sera were diluted at 1:5, and CSF samples were diluted at 1:2. Live human embryonic kidney 293 (HEK293) cells were transiently transfected with the human AQP4 M1 isoform and a non-linked green fluorescent protein (GFP) using the pIRES2-AcGFP-hAQP4/M1 plasmid. After 36 hours, both transfected AQP4-expressing (GFP-positive) and non-transfected (GFP-negative) cells were harvested and incubated with serum or CSF. Bound human IgG was detected using an Alexa Fluor 647-conjugated goat anti-human IgG (Fc-specific) secondary antibody. The presence of AQP4-IgG was quantified using the IgG binding index (IBI), calculated as the ratio of median fluorescence intensity (MFI) of transfected (GFP-positive) cells to non-transfected (GFP-negative) cells. An IBI ≥ 2.0 was considered positive.^18,19^ The ultimate diagnosis of NMOSD was made in accordance with the 2015 diagnostic criteria, in cases with anti-AQP4 antibodies.^20^

To measure MOG-IgG, HEK293 cells were transiently transfected with a DNA plasmid co-expressing full-length human MOG and AcGFP. After 36 hours, both transinfected MOG-expressing (GFP-positive) and non-transinfected (GFP-negative) cells were harvested and incubated with serum or CSF. After washing, Alexa Fluor 647-conjugated anti-human IgG1 secondary antibody was applied for serum and Alexa Fluor 647-conjugated goat anti-human IgG (Fc-specific) secondary antibody for CSF. Cells were analyzed by flow cytometry, gating on GFP-positive and GFP-negative populations. The MOG-IgG binding index was calculated as the ratio of MFI in GFP-positive cells to GFP-negative cells, with values >2·5 considered positive in both serum and CSF as previously described.^21,22^ MOGAD was ultimately diagnosed based on the established diagnostic criteria.^23,24^

### Statistical Analysis

Categorical variables such as sex at birth, autoimmune disease status, clinical presentation, neurological exam findings, MRI features, and CSF parameters were summarized as frequencies and proportions while continuous variables were reported as medians and interquartile ranges (IQR). Analyses were stratified by autoimmune status, mNGS result, vitamin B12 level, presence of tumor, vascular malformation, and HIV status. All analyses were conducted using Stata version 18 (StataCorp, College Station, TX)

## Results

We evaluated 420 participants with suspected non-traumatic myelopathy (**Figure 1**). Of these, 219 were excluded: 150 due to extradural compressive lesions on MRI, eight with alternative diagnoses, and 61 who declined consent, mainly due to fear of lumbar puncture. Among the 201 individuals with non-compressive myelopathy, 57 were excluded for lacking both CSF and serum diagnostic studies, while inclusion required having serum and/or CSF testing. The final analysis included 144 individuals with non-compressive myelopathy. Characteristic MRI findings were used to establish the etiology of non-compressive myelopathy (**Figure 2**).

**Figure 1:**
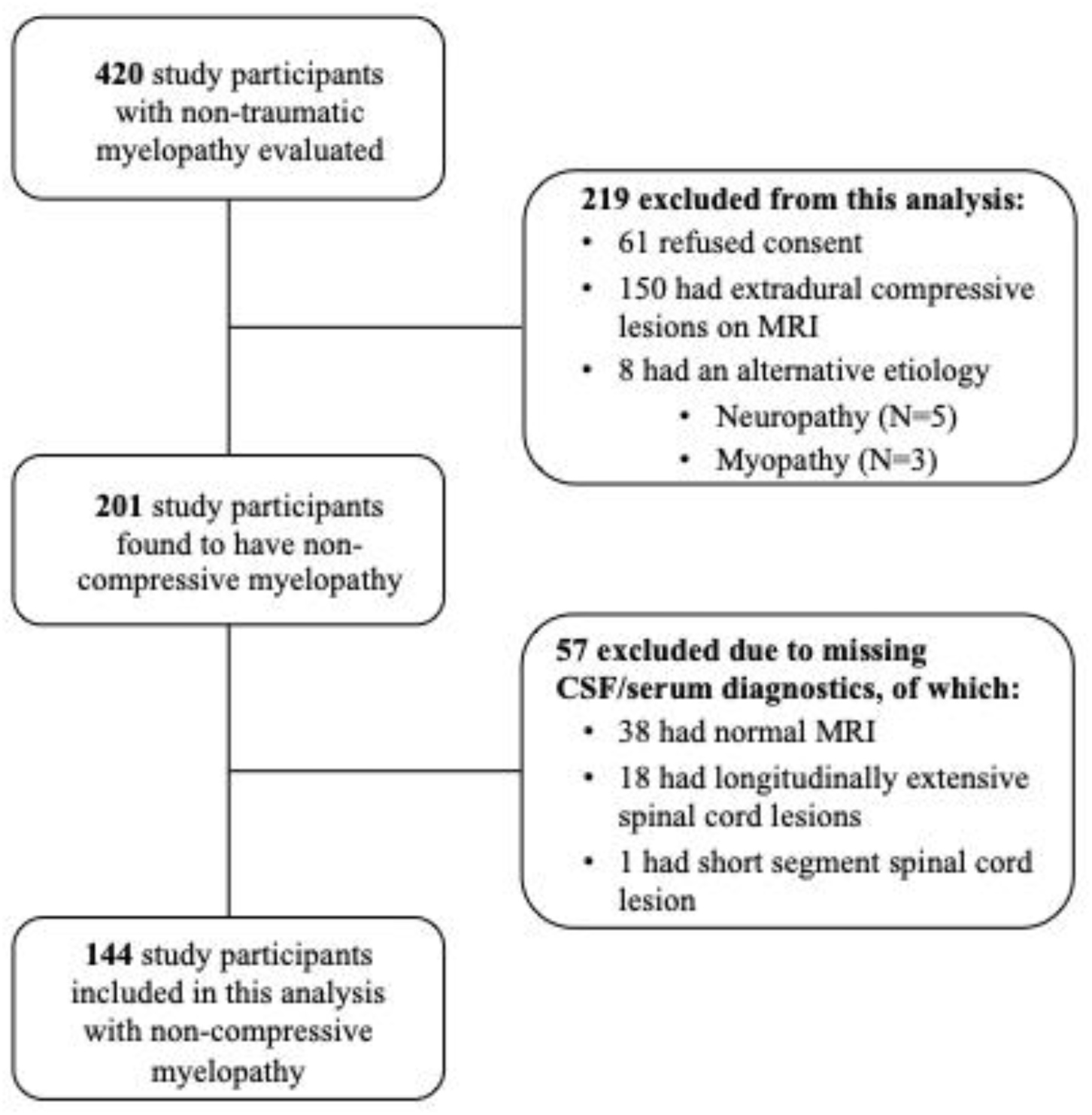
Participant Flow Diagram Showing Inclusion of Ugandan Participants with Non-compressive Myelopathy.

**Figure 2:**
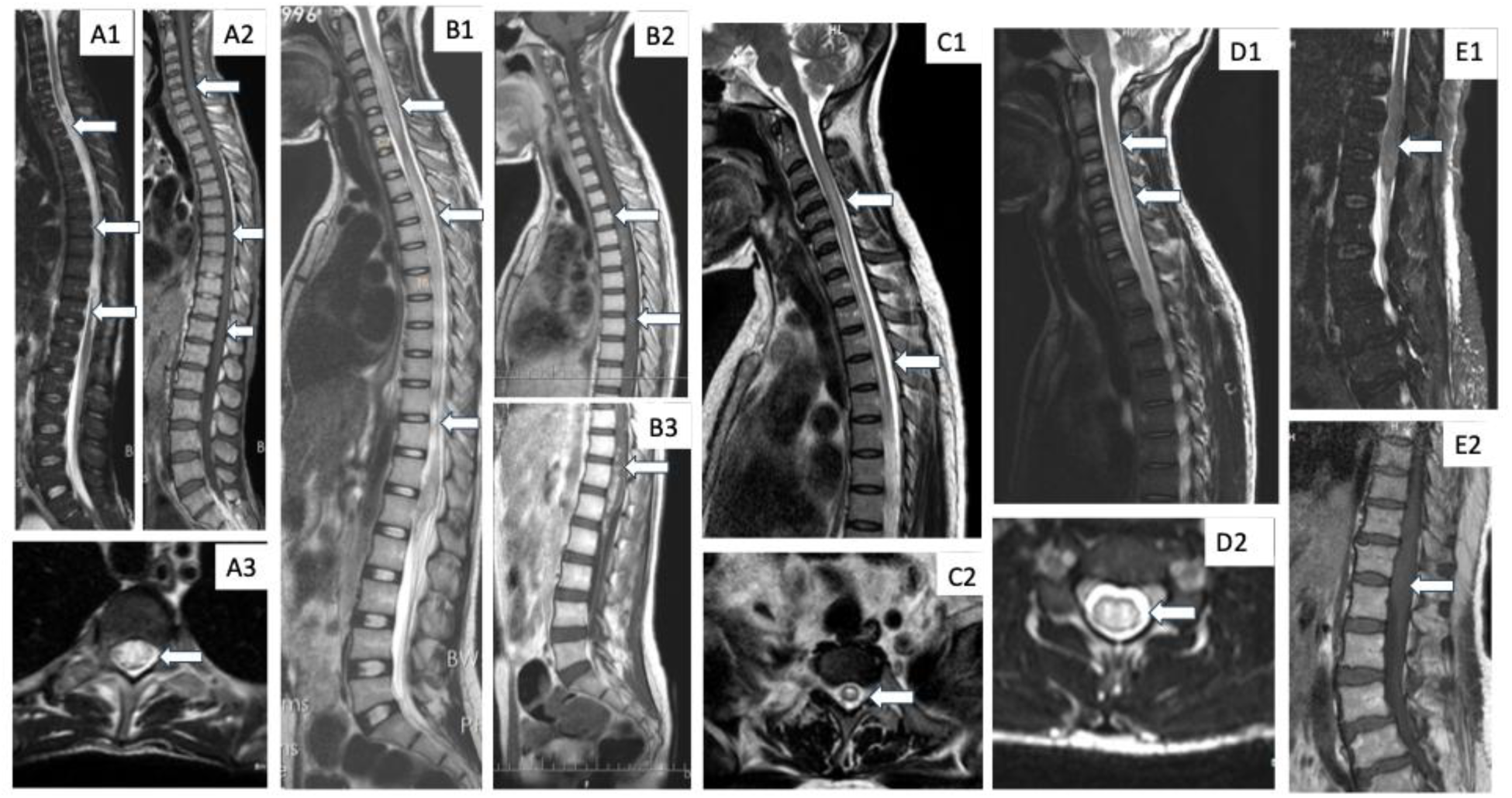
Representative MRI Findings in Ugandan Participants with Confirmed Etiologies of Non-Compressive Myelopathy A: Varicella zoster virus (VZV) – A1) Sagittal T2-weighted MRI shows a longitudinally extensive spinal cord lesion spanning the cervical and thoracic regions. A2) Sagittal post-contrast T1-weighted image demonstrates enhancement in the cervical spinal cord. A3) Axial T2-weighted image reveals involvement of more than two-thirds of the spinal cord cross-sectional area. **B: Neuroschistosomiasis** – B1) Sagittal T2-weighted MRI shows a longitudinally extensive lesion involving the entire length of the spinal cord. B2) Sagittal post-contrast T1-weighted image demonstrates patchy enhancement in the cervical and thoracic spinal cord. B3) Additional post-contrast enhancement is seen in the lumbar spine, primarily involving the conus medullaris. **C: Neuromyelitis Optica spectrum disorders** – C1) Sagittal T2-weighted MRI shows a longitudinally extensive lesion in the cervical and thoracic spinal cord. C2) Axial T2-weighted image demonstrates a centrally located hyperintense lesion. **D: Myelin oligodendrocyte glycoprotein antibody disease** – D1) Sagittal T2-weighted MRI shows a longitudinally extensive lesion in the cervical spinal cord. D2) Axial T2-weighted image reveals a centrally located hyperintense lesion. **E: Germ cell tumor confirmed on a biopsy** - E1) Sagittal T2-weighted MRI shows an expansile, mildly hyperintense lesion at the conus medullaris. E2) post-contrast sagittal T1-weighted image shows no contrast enhancement.

Of the 144 individuals with non-compressive myelopathy, 26% (n=38) were living with HIV, 55% (n=79) were male, and the median age was 33 years [IQR 25 – 44] (**Table 1**). The most common presenting signs and symptoms were abnormal sensation (69%, n=100), bladder or bowel dysfunction (65%, n=94), and paraparesis or paraplegia (55%, n=79), with the majority (76%, n=110) presenting with two or more symptoms. Severe disease was observed in 47% of participants, based on the ASIA Impairment Scale Grade A (n=42) or Grade B (n=26). Abnormal MRI spine findings were present in 55% (n=79/144), of whom 85% (n=67/79) had a long-segment, T2-weighted hyperintense lesion. CSF analysis was performed in 93 of the 144 participants enrolled in the study. Elevated protein was present in 63% (n=59/93), and pleocytosis (white cell count >5 cells/mm³) was observed in 30% (n=28/93). CSF Gram stain and culture were negative in all cases.

**Table 1:** Baseline Characteristics of Ugandan Adults Presenting with Non-Compressive Myelopathy.

| Characteristics | N | N (%)<br>Median [IQR] |
| --- | --- | --- |
| **Total, No.** | 144 |  |
| **Demographics** |  |  |
| Sex, male | 144 | 79 (55%) |
| Age, years | 144 | 33 [25 - 44] |
| <25 |  | 29 (20%) |
| 25 – 49 |  | 92 (64%) |
| >50 |  | 23 (16%) |
| HIV, seropositive | 144 | 38 (26%) |
| **Clinical History** |  |  |
| Symptoms duration, weeks | 144 | 27 [11 - 113] |
| <2 |  | 52 (26%) |
| 2 – 4 |  | 23 (16%) |
| >4 |  | 69 (48%) |
| HIV, seropositive | 144 | 38 (26%) |
| **Signs and Symptoms** | 144 |  |
| Vision abnormalities |  | 3 (2%) |
| Encephalopathy |  | 6 (4%) |
| Abnormal sensation |  | 100 (69%) |
| Quadriparesis/Quadriplegia |  | 30 (21%) |
| Paraparesis/Paraplegia |  | 79 (55%) |
| Bladder/Bowel dysfunction |  | 94 (65%) |
| **Number of Symptoms** | 144 |  |
| >2 |  | 110 (76%) |
| >3 |  | 31 (22%) |
| >4 |  | 3 (2%) |
| **American Spinal Injury Association (ASIA) Scale** | 144 |  |
| Grade A |  | 42 (29%) |
| Grade B |  | 26 (18%) |
| Grade C |  | 46 (32%) |
| Grade D |  | 30 (20%) |
| **Cerebrospinal Fluid Characteristics** |  |  |
| Protein >45 mg/dL | 93 | 59 (63%) |
| White blood cell >5 cells/μL | 93 | 28 (30%) |
| **MRI Characteristics** | 144 |  |
| Normal spinal cord |  | 65 (45%) |
| Abnormal spinal cord<sup>1</sup> |  | 79 (55%) |
| Spinal cord atrophy |  | 8 (10%) |
| Short segment T2W hyperintense lesions |  | 4 (5%) |
| Long segment T2W hyperintense lesions<sup>2</sup> |  | 67 (85%) |
| With conus medullaris involvement |  | 7 (10%) |
| Without conus medullaris involvement |  | 36 (54%) |
| Posterior location |  | 4 (6%) |
| Prominent flow voids of the spinal cord |  | 2 (3%) |
| With Leptomeningeal involvement |  | 6 (9%) |
| With Spinal cord expansion |  | 12 (18%) |

<sup>1</sup>Abnormal MRI images were further characterized by spinal cord morphology and lesion type.
<sup>2</sup>Long-segment T2-weighted hyperintense lesions were further categorized by anatomical and radiological features.

CSF mNGS was performed on stored samples from 77 study participants, identifying pathogens in 13 (17%) cases (**Table 2**). Among these, *Schistosoma mansoni* was detected in eight individuals, cytomegalovirus (CMV) in one, and varicella-zoster virus (VZV) in one. HIV-1 was identified as the sole pathogen in two individuals and co-detected in the CMV and VZV cases. A heatmap of the detected pathogens and corresponding reads per million (combined RNA/DNA) is shown in **Figure 3**. *S. mansoni*, VZV, and CMV were presumed to be causative pathogens in their respective cases.

**Table 2:** Cerebrospinal fluid and Serum Diagnostic Testing in Ugandan Adults with Non-Compressive Myelopathy.

| Diagnostic Test | N | Positive N (%) |
| --- | --- | --- |
| **Metagenomic next generation sequencing, CSF** | 77 |  |
| *Schistosoma mansoni* |  | 8 (10%) |
| HIV |  | 2 (3%) |
| VZV/HIV* |  | 1 (1%) |
| CMV/HIV* |  | 1 (1%) |
| *Cryptococcus Neoformans* |  | 1 (1%) |
| **Antibodies, CSF/Serum** | 121 |  |
| Aquaporin-4 immunoglobulin (AQP4-IgG) |  | 10 (8%) |
| Myelin oligodendrocyte glycoprotein-immunoglobulin G (MOG IgG) |  | 10 (8%) |
| **Vitamin B12, Serum** | 22 |  |
| ≤ 300 pg/mL |  | 9 (41%) |
| > 300 pg/mL |  | 13 (59%) |
| Abbreviations: VZV; Varicella-zoster virus, CMV; Cytomegalovirus<br>*More than one organism was detected on metagenomic next-generation sequencing in the same individual |  |  |

**Figure 3:**
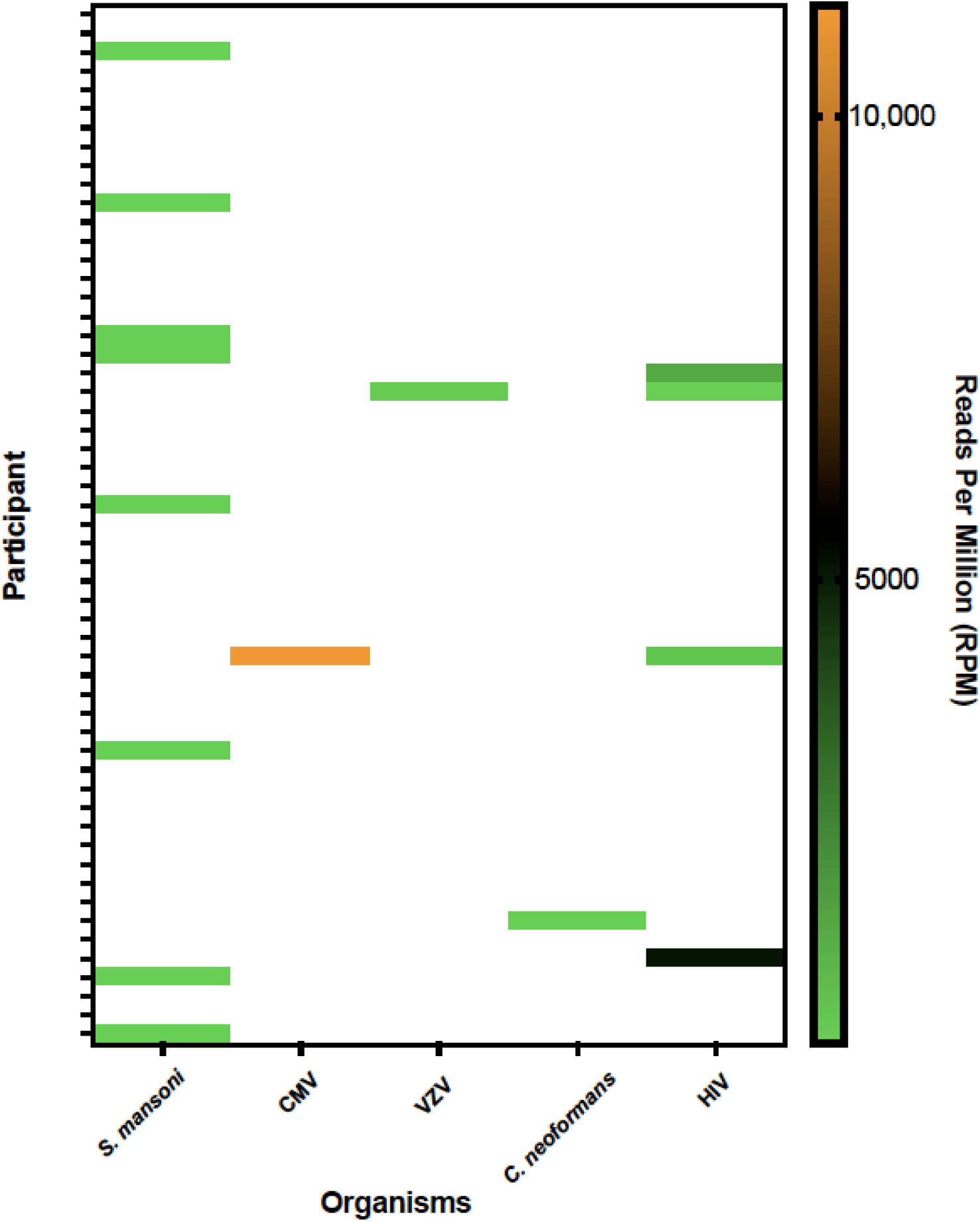
Metagenomic Next Generation Sequencing Read Depth Profiles of Ugandan Participants with non-compressive Myelopathy. Heatmap of participants sequenced, and organisms identified on metagenomic next generation sequencing. The color map and scale quantify the reads per million (RPM) from either the ribonucleic acid (RNA) or deoxyribonucleic acid (DNA) sequencing of the organism identified per sample. Five organisms were identified including: *Schistosoma mansoni*, Cytomegalovirus (CMV), Varicella Zoster Virus (VZV), Human Immunodeficiency Virus (HIV) and *Cryptococcal neoformans* In the participants with CMV and VZV identified with high RPMs, there was co-infection with HIV in both.

Confirmatory *S. mansoni* PCR was positive in all mNGS *S. mansoni* positive cases.^25^ Ultimately, we identified nine cases of neuroschistosomiasis: eight were diagnosed by mNGS and one by spinal cord biopsy (**Table 4**). Two individuals (22%) were female, none were HIV-positive, and the median time from symptom onset to presentation was 8 days [IQR 6 – 22]. Three participants (33%) presented with severe neurological impairment (ASIA grade A or B), and four (44%) had CSF pleocytosis. All nine presented with long-segment T2-weighted hyperintense spinal cord lesions.

In the two individuals with HIV-1 alone, the etiologic role of HIV-1 was uncertain. In both patients, the MRI findings were atypical for HIV-associated vacuolar myelopathy: one exhibited a central longitudinally extensive transverse myelitis (LETM) lesion within 60 days of symptom onset at a CD4 count of 90 cells/mm^3^, whereas the other showed disease progression over 113 days despite a normal MRI. Based on the clinical presentation, detection of *Cryptococcus neoformans* on mNGS was not considered a likely contributor to the observed myelopathy in one study participant. Human pegivirus type 1 was detected in 6 cases with unclear clinical significance^26,27^.

Serum and CSF antibody testing for AQP4-IgG and MOG-IgG was performed in 121 study participants, with 10 testing positive for AQP4-IgG and 10 for MOG-IgG (**Tables 2 and3**). The majority of AQP4-IgG IBI levels were high in both serum and CSF, whereas MOG-IgG IBI levels were generally low. Following clinical adjudication, nine AQP4-IgG–positive individuals fulfilled criteria for definite NMOSD and one for possible NMOSD. Six MOG-IgG–positive individuals met criteria for definite MOGAD and one for possible MOGAD.

**Table 3:** Serum and Cerebrospinal Fluid Aquaporin-4 and Myelin Oligodendrocyte Glycoprotein Antibody Positivity in Ugandan Participants with Non-Compressive Myelopathy.

| Aquaporin-4-IgG |  |  |  |  |  |
| --- | --- | --- | --- | --- | --- |
| Participant | Time to Presentation<sup>1</sup> | Binding Index<sup>2</sup>, Serum | Binding Index<sup>2</sup>, CSF | MRI Findings | Interpretation<sup>4</sup> |
| 1. | 20 | 63.51 | 30.38 | LETM | definite NMOSD |
| 2. | 14 | 24.26 | 2.3 | LETM | definite NMOSD |
| 3. | 5 | 24.21 | 53.31 | LETM | definite NMOSD |
| 4. | 27 | 19.2 | 64.92 | ATROPHY** | Definite NMOSD |
| 5. | 15 | 18.41 | 1.57 | LETM | definite NMOSD |
| 6. | 124 | 16.04 | 4.71 | LETM | definite NMOSD |
| 7. | 86 | 11.69 | 1.41 | OPTIC NEURITIS | definite NMOSD |
| 8. | 366 | 3.43 | 2 | LETM | definite NMOSD |
| 9. | 3 | 2.25 | - | NORMAL** | Possible NMOSD |
| 10. | 6 | 0.89 | 15.37 | ATROPHY | Definite NMOSD |
| Myelin Oligodendrocyte Glycoprotein-IgG |  |  |  |  |  |
| Participant | Time to Presentation<sup>1</sup> | Binding Index<sup>3</sup>, Serum | Binding Index<sup>3</sup>, CSF |  | Interpretation<sup>4</sup> |
| 1. | 37 | 3.88 | 1.07 | normal* | Possible MOGAD |
| 2. | 16 | 3.06 | 1.69 | TUMOR | False Positive test |
| 3. | 17 | 2.83 | 3.92 | STM* | Definite MOGAD |
| 4. | 60 | 2.59 | 3.26 | TUMOR | False Positive test |
| 5. | 42 | 2.53 | 2.7 | LETM | Definite MOGAD |
| 6. | 1 | 2.53 | - | LETM | Definite MOGAD |
| 7. | 33 | 1.68 | 3.39 | STM* | Definite MOGAD |
| 8. | 6 | 1.59 | 2.74 | TUMOR | False Positive test |
| 9. | 12 | 1.25 | 3.01 | LETM | Definite MOGAD |
| 10. | 2 | 1.22 | 3.77 | LETM | Definite MOGAD |
| <sup>1</sup>Time to presentation is reported in days<br><sup>2</sup>An IgG binding index of $\geq 2.0$ is considered a positive result, indicating the presence of AQP4-IgG antibodies.<br><sup>3</sup>An IgG binding index of $\geq 2.5$ is considered a positive result, indicating the presence of MOG-IgG antibodies.<br><sup>4</sup>True positive and false positives were determined based on positive antibody response and clinical adjudication using the NMOSD and MOGAD diagnostic criteria.<br>LETM-Longitudinally Extensive Transverse Myelitis, STM- Short Transverse Myelitis<br>Atrophy and Normal MRI findings were regarded as possible NMOSD<br>Short transverse myelitis lesions were also included as definite MOGAD disease because 20% are present with such lesions and 10% present with a normal MRI<br>Patient Number 9 : MRI normal. This could be early disease | | | | | |

**Table 4:** Baseline Characteristics and One-Year Mortality Based by Etiological Diagnosis of Non-Compressive Myelopathy in Uganda Participants.

| Characteristics | NMOSD | MOGAD | Neuroschistosomiasis | Vitamin B12 Myelopathy | Tumor |
| --- | --- | --- | --- | --- | --- |
| No. | 10 | 7 | 9 | 8 | 8 |
| Age, years | 42 [31 – 48] | 25 [23 – 36] | 26 [23 – 27] | 31 [25 – 38] | 32 [40 – 63] |
| Sex, female | 8 (80%) | 1 (14%) | 2 (22%) | 3 (38%) | 5 (63%) |
| HIV-serostatus, positive | 1 (10%) | 2 (29%) | 0 (0%) | 1 (13%) | 0(0%) |
| **Clinical Presentation** |  |  |  |  |  |
| Disease duration, days | 18 [8 – 71] | 17 [7 – 35] | 8 [6 – 22] | 157 [46 – 698] | 173[49 -423] |
| ASIA Impairment Scale, A or B | 5 (50%) | 5 (71%) | 3 (33%) | 1 (13%) | 4 (50%) |
| CSF white cell count >5 cells/μL | 0 (0%) | 2/6** (33%) | 4 (44%) | 0* | 0 (0%) *** |
| **MRI Characteristics** |  |  |  |  |  |
| Optic neuritis | 1 | 0 | 0 | 0 | 0 |
| Normal spinal Cord | 1 | 1 | 0 | 7 | 0 |
| Spinal cord atrophy | 2 |  |  |  |  |
| Short segment T2W hyperintense lesions |  | 2 |  |  |  |
| Abnormal spinal cord | 6 | 4 | 9 | 1 | 12 |
| Long segment T2W hyperintense lesions | 6 | 4 | 9 | 0 | 0 |
| Posterior Location | 0 | 0 | 0 | 1 | 0 |
| With spinal cord expansion | 0 | 0 | 0 | 0 | 8 |
| All-cause one-year mortality | 4 (40%) | 2 (29%) | 1 (11%) | 1 (13%) | 4 (50%) |

**Abbreviations:** ASIA, American Spinal Injury Association; NMOSD, Neuromyelitis Optica Spectrum Disorder; MOGAD, Myelin Oligodendrocyte Glycoprotein Antibody-Associated Disease.
Data is presented as either N(%) or median [IQR]
\*CSF white cell count analysis was performed for one participant.
\*\*CSF white cell count analysis was performed for six participants.
\*\*\*CSF white cell count analysis was performed in four participants

Individuals with NMOSD and MOGAD generally presented within the first month of disease onset and with severe manifestations. Among the NMOSD cases, one patient developed optic neuritis four months after an episode of paraparesis despite a normal spinal MRI, while brain MRI confirmed optic neuritis; two others demonstrated spinal cord atrophy on MRI. Most NMOSD patients were female (80%; n=8), and one was co-infected with HIV. In contrast, none of the seven individuals with MOGAD were female, and two were co-infected with HIV. The possible NMOSD case had imaging performed three days after disease onset, whereas the possible MOGAD case had a normal MRI 37 days after onset. Notably, three MOG-IgG– positive individuals who did not fulfill diagnostic criteria for MOGAD demonstrated expansile imaging lesions more suggestive of neoplasm than demyelinating disease (Table 4).

The most common etiology of non-inflammatory myelopathy was vitamin B12 deficiency. Serum vitamin B12 levels were measured in only 22 participants, nine of whom had a serum level ≤ 300 pg/mL (**Table 2**). One individual with a low serum vitamin B12 level of 37 pg/mL tested positive for MOG-IgG and had a longitudinally extensive T2 hyperintense spinal cord lesion on MRI; this case was classified as MOGAD due to the longitudinally extensive lesion and the short disease duration– an acute onset of 12 days. Among the remaining eight individuals with low vitamin B12 levels, one exhibited a longitudinal, posterior column T2 signal hyperintensity on spinal MRI, a finding typically associated with subacute combined degeneration from vitamin B12 deficiency (**Table 4**). The other seven individuals had normal spinal MRIs. The eight individuals with vitamin B12 myelopathy had the following clinical characteristics: three were female, one had HIV co-infection, and time from symptom onset to presentation varied widely (median 157 days [IQR 46–698]), reflective of the more chronic, progressive course of subacute combined degeneration. Disease severity was generally mild, with only one (13%) individual presenting with severe deficits, and one death (13%) reported at one-year follow-up.

Among the 12 patients with spinal cord expansion, four were considered to have granulomatous disease, but we could not determine a specific diagnosis in these cases. The remaining eight were diagnosed with spinal cord tumors: seven were identified through expansile MRI lesions, and one was biopsy-confirmed (Table 4). Of these, one patient had a lung mass, two had infiltrative masses, and 4 showed nodular enhancement lesions consistent with possible metastatic disease. Median time to the presentation was 173 days.

The two vascular cases demonstrated flow voids on MRI suggestive of arteriovenous malformation (AVM). Both patients were young (18 and 28, one male, one female), and disease onset was 14 days and 166 days, respectively. The participant with delayed presentation of 166 days from onset died after one year.

Of the 144 participants enrolled in this observational study, we established a definitive etiologic diagnosis in 46 individuals (32%) (**Table 5)**. Most of those with a confirmed diagnosis (n=36/46, had abnormal spinal cord MRI findings. Among the 65 participants (45%) with a normal MRI, ten received a confirmed diagnosis—primarily vitamin B12 deficiency (n=7). We were unable to make a definitive diagnosis in 98 (68%) individuals who presented with signs and symptoms suggestive of non-compressive myelopathy.

**Table 4:**
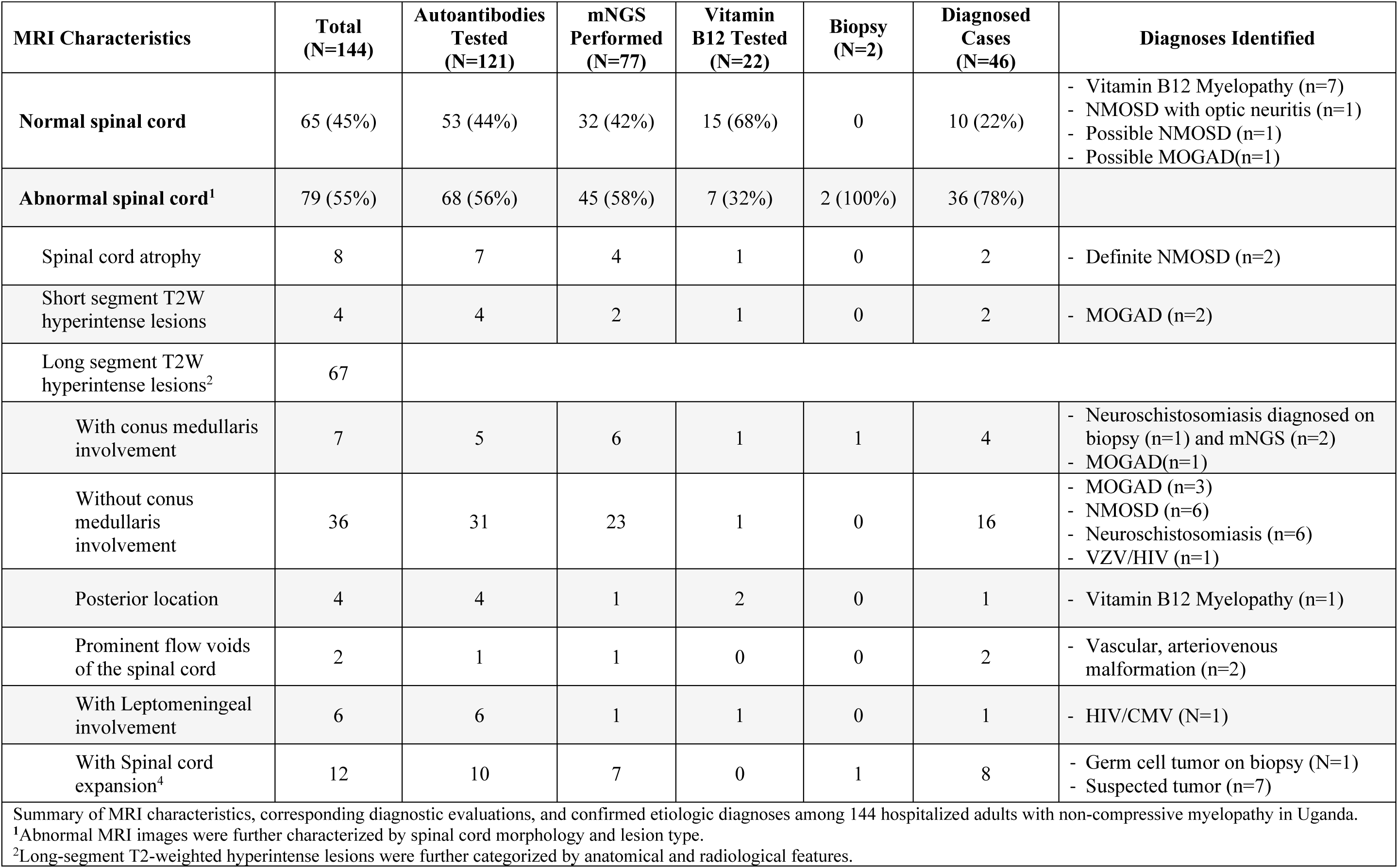
MRI Characteristics, Diagnostic Testing, and Confirmed Etiologies of Non-Compressive Myelopathy Among Hospitalized Ugandan Adults.

## Discussion

To our knowledge, this is the first study to comprehensively investigate the etiologies of non-compressive myelopathy in a hospitalized population in Uganda using advanced diagnostic tools. The use of mNGS enabled identification of an infectious etiology in 10 individuals, yielding a diagnostic rate of 13% among those tested. Autoantibody testing identified 17 cases of autoimmune-mediated myelopathy, yielding a 14% diagnostic yield among specimens tested. Vitamin B12 testing was particularly informative in individuals with normal spinal MRI; among the 22 individuals tested, eight (36%) were diagnosed with B12-associated myelopathy.

Furthermore, several participants exhibited imaging features suggestive of AVM or spinal tumors, with one case subsequently confirmed through biopsy. Despite the extensive diagnostic workup, an etiologic diagnosis could not be established in 68% of participants, highlighting the continued diagnostic challenges in this population.

The prevalence and causes of non-compressive myelopathy remain poorly understood across Africa. Most prior studies in the region have focused on compressive spinal cord lesions and lacked systemic evaluation for on non-compressive myelopathies.^4^ In a study of 58 Malawians with nontraumatic paraplegia of less than six months duration, a definitive diagnosis was made in 80% of cases, most commonly spinal tuberculosis, tumors, and TM, while 10% remained undiagnosed. The retrospective use of real-time PCR further identified infections such as *Treponema pallidum*, *S. mansoni*, Epstein-Barr virus, and herpes simplex virus 2 as causes of TM.^9^ In another study by Modi et al. involving 100 individuals, 50% of whom were living with HIV, pathogen-specific PCR and other diagnostic testing identified several causes of non-compressive myelopathy including: HIV-associated vacuolar myelopathy (n=8), CMV (n=1), VZV (n=2), acute disseminated encephalomyelitis (n=4), NMOSD (n=2), and B12 deficiency (n=1).^10^

In our study, we identified a similar number of myelopathy cases due to infectious and autoimmune causes, highlighting the importance of incorporating autoimmune testing in African settings. NMOSD is the most extensively studied CNS autoimmune condition in Africa, with most studies conducted in North Africa and only a few in Sub-Saharan Africa.^8,28–30^ In contrast, MOGAD remains largely unexplored in African populations, with only a single published case report.^31^ We used a live cell-based assay for AQP4-IgG testing that has the highest sensitivity (94%) and specificity (>99.5%).^32^ Using this approach, we classified spinal cord atrophy as definite NMOSD, while a normal MRI was considered possible NMOSD, consistent with early disease presentation. Other demyelinating phenotypes, including optic neuritis, demyelinating disease of the brain, and AQP4-negative NMOSD were not systematically captured by this study.^33^ MOGAD was considered definite in patients with short TM lesions, consistent with reports that this occurs in approximately 15–30% of patients.^24,34^ A possible MOGAD diagnosis was assigned to a patient with a normal MRI, reported in about 10% of cases.^24,35^ Missed diagnoses are most likely in MOGAD, as TM occurs in only one-third of patients, while 70% present with optic neuritis or demyelination in the brain, which were not systematically captured in this study.^36^ The MOG-IgG assay demonstrates a sensitivity of 75–92% and specificity of 93– 100%.^21^ ^37^

CSF mNGS is a powerful tool for diagnosing neurologic infections.^38^ In settings with a high burden of infectious diseases and a high prevalence of advanced HIV, there is a critical need for unbiased diagnostic approaches.^4^ In addition to VZV and CMV, we detected 8 cases of neuroschistosomiasis through mNGS (9 total). The true prevalence of neuroschistosomiasis in Uganda remains unknown; however, it is likely to be substantial given the high national burden of schistosomiasis, estimated at 25.6%^39^ Notably, neuroschistosomiasis is a treatable condition, with both anti-parasitic and anti-inflammatory therapies available within the Ugandan healthcare system. Expanding routine screening for *Schistosoma mansoni* may therefore facilitate earlier recognition of neurological involvement and lead to improved clinical outcomes for affected individuals.

We also found vitamin B12 deficiency to be one of the most common non-inflammatory etiologies of myelopathy despite limited testing in our cohort. This finding is consistent with two other studies that have reported low vitamin B12 levels in African populations.^10,40^ Lastly, imaging also enabled the diagnosis of spinal cord tumors and vascular causes of myelopathy in a subset of patients.

### Limitations

This study had several limitations. Due to financial constraints, neuroimaging was focused exclusively on the spinal cord, potentially missing diagnoses such as MS, which often requires the identification of demyelinating lesions in the brain and spinal cord. In some cases, MRI was performed without gadolinium contrast, limiting the ability to detect active inflammation or enhancing lesions suggestive of specific etiologies. Spinal angiography was not performed due to lack of availability in Uganda, which limited evaluation for vascular causes of myelopathy. We were unable to assess for intrathecal immunoglobulin production, such as oligoclonal bands or kappa free light chains. Visual assessments were not performed systematically. Other metabolic causes, such as copper deficiency, were not evaluated. Additionally, we did not assess spinal cord sarcoidosis, connective tissue diseases (e.g., systemic lupus erythematosus) or genetic causes of myelopathy, which represent important avenues for future investigation.

### Conclusions

Using a combination of clinical phenotyping, neuroimaging, autoantibody testing, CSF mNGS, and limited metabolic evaluation, we identified a definitive etiology for non-compressive myelopathy in 46 (32%) participants, representing the largest prospective study of its kind in Sub-Saharan Africa. Many of the diagnoses were treatable, including NMOSD, MOGAD, infectious myelopathies, vitamin B12 deficiency, vascular malformations, and spinal cord tumors. In addition to neuroimaging, incorporating mNGS for pathogen detection, and expanding access to AQP4-IgG and MOG-IgG testing can further improve diagnostic yield and potentially lead to better clinical outcomes for individuals with non-compressive myelopathy in Uganda and similar settings across Africa.

## Contributors

Conceptualization: [Name(s)] Abdu K Musubire, David B Meya Paul R Bohjanen, Sean J. Pittock, David R Boulware, Michael R. Wilson, Patrick Cras

Methodology: [Name(s)] Abdu K Musubire, Kristoffer E Leon, Prashanth S Ramachandran, Sean J. Pittock, Eoin P Flanagan, Jospeph L. DeRisi, Michael R. Wilson

Formal analysis and investigation: [Name(s)] Abdu K Musubire Kristoffer E Leon, Prashanth S Ramachandran, Chloe Gerungan, Kelsey C Zorn, Annie Wapniarski, Paul Mubiri M, Twaha Kisozi, Eoin P Flanagan, Kimbugwe Denis, Mahsa Abassi, Jessica Sagen, Barbara Willekens

Data curation: [Name(s)] Abdu K Musubire Kristoffer E Leon, Prashanth S Ramachandran, Cynthia T Chin Barbara Willekens, Eoin P Flanagan, Mahsa Abassi

Writing – original draft preparation: [Name(s)] Abdu K Musubire, Kristoffer E Leon, Mahsa Abassi, Michael R. Wilson, Barbara Willekens

Writing – review and editing: [Name(s)] Michael R. Wilson Barbara Willekens, Eoin P Flanagan, Kristoffer E Leon, Prashanth S Ramachandran, Cynthia T Chin, Kelsey C Zorn, Annie Wapniarski, David B Meya, Paul Mubiri M, Twaha Kisozi, Mahsa Abassi, David R Boulware, Vyanka Redenbaugh, Jessica Sagen, Paul R Bohjanen, Barbara Willekens, Sean J. Pittock

Supervision: [Name(s)] David B Meya Paul R Bohjanen, Sean J. Pittock Michael R. Wilson Barbara Willekens, David R Boulware, Patrick Cras

Funding acquisition: [Name(s)] Abdu K Musubire, Paul R Bohjanen, Michael R. Wilson, David R Boulware

All authors reviewed, revised, and approved the final version of the manuscript.

## Data sharing

Deidentified participant data supporting the findings of this article will be made accessible to qualified investigators upon reasonable request to the corresponding author between 12- and 36-months following publication. Non-host sequences were deposited in the NCBI Sequence Read Archive (SRA) with the primary accession code PRJNA1270729

## Data Availability

All data produced in the present study are available upon reasonable request to the authors

## Acknowledgements

I would like to acknowledge the contributions of the following people: Prof Kawooya, Prof Carlos Pardo, Dr. Twaha Kisozi, Dr. Henry Nabeta, Dr. Matovu Stephen, Dr. Edward Mpoza, Cynthia Ahimbisibwe, Jane Buyonjo. We thank the patients and their families for participation in this study.

## References

1. Blackburn KM, Greenberg BM. Revisiting Transverse Myelitis: Moving Toward a New Nomenclature. Frontiers in neurology 2020; 11.

2. Malekzadeh H, Golpayegani M, Ghodsi Z, et al. Direct Cost of Illness for Spinal Cord Injury: A Systematic Review. Global spine journal 2022; 12(6): 1267–81.

3. New PW, Cripps RA, Bonne Lee B. Global maps of non-traumatic spinal cord injury epidemiology: towards a living data repository. Spinal cord 2014; 52(2): 97–109.

4. Musubire AK, Meya DB, Bohjanen PR, et al. A Systematic Review of Non-Traumatic Spinal Cord Injuries in Sub-Saharan Africa and a Proposed Diagnostic Algorithm for Resource-Limited Settings. Frontiers in neurology 2017; 8: 618.

5. Carnero Contentti E, Hryb JP, Diego A, Di Pace JL, Perassolo M. Etiologic spectrum and functional outcome of the acute inflammatory myelitis. Acta neurologica Belgica 2017; 117(2): 507–13.

6. Gastaldi M, Marchioni E, Banfi P, et al. Predictors of outcome in a large retrospective cohort of patients with transverse myelitis. Multiple sclerosis (Houndmills, Basingstoke, England) 2018; 24(13): 1743–52.

7. Alkabie S, Casserly CS, Morrow SA, Racosta JM. Identifying specific myelopathy etiologies in the evaluation of suspected myelitis: A retrospective analysis. Journal of the neurological sciences 2023; 450: 120677.

8. Musubire AK, Derdelinckx J, Reynders T, et al. Neuromyelitis Optica Spectrum Disorders in Africa: A Systematic Review. Neurology(R) neuroimmunology & neuroinflammation 2021; 8(6).

9. Zijlstra EE, van Hellemond JJ, Moes AD, et al. Nontraumatic Myelopathy in Malawi: A Prospective Study in an Area with High HIV Prevalence. The American journal of tropical medicine and hygiene 2020; 102(2): 451–7.

10. Modi G, Ranchhod J, Hari K, Mochan A, Modi M. Non-traumatic myelopathy at the Chris Hani Baragwanath Hospital, South Africa--the influence of HIV. QJM : monthly journal of the Association of Physicians 2011; 104(8): 697–703.

11. Sechi E, Shosha E, Williams JP, et al. Aquaporin-4 and MOG autoantibody discovery in idiopathic transverse myelitis epidemiology. Neurology 2019; 93(4): e414–e20.

12. Tremlett H, Chomba M, Mortel D, et al. Comorbidity in the multiple sclerosis and neuromyelitis optica spectrum disorders population: findings from an underserved, low income country, Zambia. Multiple sclerosis and related disorders 2024; 81: 105365.

13. Jamal I, Shah J, Mativo P, Hooker J, Wallin M, Sokhi DS. Multiple sclerosis in Kenya: Demographic and clinical characteristics of a registry cohort. Multiple sclerosis journal - experimental, translational and clinical 2021; 7(2): 20552173211022782.

14. Musubire AK, Meya DB, Katabira ET, et al. Epidemiology of non-traumatic spinal cord injury in Uganda: a single center, prospective study with MRI evaluation. BMC neurology 2019; 19(1): 10.

15. Linassi G, Li Pi Shan R, Marino RJ. A web-based computer program to determine the ASIA impairment classification. Spinal cord 2010; 48(2): 100–4.

16. Ramachandran PS, Ramesh A, Creswell FV, et al. Integrating central nervous system metagenomics and host response for diagnosis of tuberculosis meningitis and its mimics. Nature communications 2022; 13(1): 1675.

17. Kalantar KL, Carvalho T, de Bourcy CFA, et al. IDseq-An open source cloud-based pipeline and analysis service for metagenomic pathogen detection and monitoring. Gigascience 2020; 9(10).

18. Redenbaugh V, Montalvo M, Sechi E, et al. Diagnostic value of aquaporin-4-IgG live cell based assay in neuromyelitis optica spectrum disorders. Multiple sclerosis journal - experimental, translational and clinical 2021; 7(4): 20552173211052656.

19. Majed M, Fryer JP, McKeon A, Lennon VA, Pittock SJ. Clinical utility of testing AQP4-IgG in CSF: Guidance for physicians. Neurology(R) neuroimmunology & neuroinflammation 2016; 3(3): e231.

20. Wingerchuk DM, Banwell B, Bennett JL, et al. International consensus diagnostic criteria for neuromyelitis optica spectrum disorders. Neurology 2015; 85(2): 177–89.

21. Redenbaugh V, Fryer JP, Cacciaguerra L, et al. Diagnostic Utility of MOG Antibody Testing in Cerebrospinal Fluid. Annals of neurology 2024; 96(1): 34–45.

22. Sechi E, Buciuc M, Pittock SJ, et al. Positive Predictive Value of Myelin Oligodendrocyte Glycoprotein Autoantibody Testing. JAMA neurology 2021; 78(6): 741–6.

23. Voase S, Robertson NP. Diagnostic criteria for MOGAD. Journal of neurology 2024; 271(6): 3690–2.

24. Banwell B, Bennett JL, Marignier R, et al. Diagnosis of myelin oligodendrocyte glycoprotein antibody-associated disease: International MOGAD Panel proposed criteria. The Lancet Neurology 2023; 22(3): 268–82.

25. Obeng BB, Aryeetey YA, de Dood CJ, et al. Application of a circulating-cathodic-antigen (CCA) strip test and real-time PCR, in comparison with microscopy, for the detection of Schistosoma haematobium in urine samples from Ghana. Ann Trop Med Parasitol 2008; 102(7): 625–33.

26. Balcom EF, Doan MAL, Branton WG, et al. Human pegivirus-1 associated leukoencephalitis: Clinical and molecular features. Ann Neurol 2018; 84(5): 781–7.

27. Scheibe F, Melchert J, Radbruch H, et al. Pegivirus-Associated Encephalomyelitis in Immunosuppressed Patients. The New England journal of medicine 2025; 392(18): 1864–6.

28. Sokhi D, Suleiman A, Manji S, Hooker J, Mativo P. Cases of neuromyelitis optica spectrum disorder from the East Africa region, highlighting challenges in diagnostics and healthcare access. eNeurologicalSci 2021; 22: 100320.

29. Ojo AS, Balogun SA, Idowu AO. Neuromyelitis optica spectrum disorder in Africa: What is the current state of knowledge? Clinical neurology and neurosurgery 2021; 206: 106709.

30. Ibrahim EAA, Gammer F, Gassoum A. Neuromyelitis optica: a pilot study of clinical presentation and status of serological biomarker AQP4 among patients admitted to a tertiary centre in NCNS, Sudan. BMC neuroscience 2020; 21(1): 9.

31. Gadama Y, Du Preez M, Carr J, et al. Myelin oligodendrocyte glycoprotein antibody-associated disease (MOGAD) and Human Immunodeficiency virus infection: dilemmas in diagnosis and management: a case series. Journal of medical case reports 2023; 17(1): 457.

32. Prain K, Woodhall M, Vincent A, et al. AQP4 Antibody Assay Sensitivity Comparison in the Era of the 2015 Diagnostic Criteria for NMOSD. Front Neurol 2019; 10: 1028.

33. Long Y, Liang J, Wu L, et al. Different Phenotypes at Onset in Neuromyelitis Optica Spectrum Disorder Patients with Aquaporin-4 Autoimmunity. Front Neurol 2017; 8: 62.

34. Varley JA, Champsas D, Prossor T, et al. Validation of the 2023 International Diagnostic Criteria for MOGAD in a Selected Cohort of Adults and Children. Neurology 2024; 103(1): e209321.

35. Sechi E, Krecke KN, Pittock SJ, et al. Frequency and characteristics of MRI-negative myelitis associated with MOG autoantibodies. Multiple sclerosis (Houndmills, Basingstoke, England) 2021; 27(2): 303–8.

36. Sechi E, Cacciaguerra L, Chen JJ, et al. Myelin Oligodendrocyte Glycoprotein Antibody-Associated Disease (MOGAD): A Review of Clinical and MRI Features, Diagnosis, and Management. Front Neurol 2022; 13: 885218.

37. Gastaldi M, Scaranzin S, Jarius S, et al. Cell-based assays for the detection of MOG antibodies: a comparative study. Journal of neurology 2020; 267(12): 3555–64.

38. Ramachandran PS, Wilson MR. Metagenomics for neurological infections - expanding our imagination. Nat Rev Neurol 2020; 16(10): 547–56.

39. Exum NG, Kibira SPS, Ssenyonga R, et al. The prevalence of schistosomiasis in Uganda: A nationally representative population estimate to inform control programs and water and sanitation interventions. PLoS Negl Trop Dis 2019; 13(8): e0007617.

40. Lekoubou Looti AZ, Kengne AP, Djientcheu Vde P, Kuate CT, Njamnshi AK. Patterns of non-traumatic myelopathies in Yaounde (Cameroon): a hospital based study. Journal of neurology, neurosurgery, and psychiatry 2010; 81(7): 768–70.

